# Internal Validation of Automated Visual Evaluation (AVE) on Smartphone Images for Cervical Cancer Screening in a Prospective Study in Zambia

**DOI:** 10.1101/2023.07.19.23292888

**Authors:** Liming Hu, Mulindi H. Mwanahamuntu, Vikrant V. Sahasrabuddhe, Caroline Barrett, Matthew P. Horning, Ishan Shah, Zohreh Laverriere, Dipayan Banik, Ye Ji, Aaron Lunda Shibemba, Samson Chisele, Mukatimui Kalima Munalula, Friday Kaunga, Francis Musonda, Evans Malyangu, Karen Milch Hariharan, Groesbeck P. Parham

## Abstract

I.

**Objectives:** Visual inspection with acetic acid (VIA) is a low-cost approach for cervical cancer screening used in most low- and middle-income countries (LMICs) but, similar to other visual tests like histopathology, is subjective and requires sustained training and quality assurance. We developed, trained, and validated an artificial-intelligence-based “Automated Visual Evaluation” (AVE) tool that can be adapted to run on smartphones to assess smartphone-captured images of the cervix and identify precancerous lesions, helping augment performance of VIA.

**Design:** Prospective study.

**Setting:** Eight public health facilities in Zambia.

**Participants:** 8,204 women aged 25-55.

**Interventions:** Cervical images captured on commonly used low-cost smartphone models were matched with key clinical information including human immunodeficiency virus (HIV) and human papillomavirus (HPV) status, plus histopathology analysis (where applicable), to develop and train an AVE algorithm and evaluate its performance for use as a primary screen and triage test for women who are HPV positive.

**Main outcome measures:** Area under the receiver operating curve (AUC); sensitivity; specificity.

**Results:** As a general population screening for cervical precancerous lesions, AVE identified cases of cervical precancerous and cancerous (CIN2+) lesions with high performance (AUC = 0.91, 95% confidence interval [CI] = 0.89 to 0.93), which translates to a sensitivity of 85% (95% CI = 81% to 90%) and specificity of 86% (95% CI = 84% to 88%) based on maximizing the Youden’s index. This represents a considerable improvement over VIA, which a meta-analysis by the World Health Organization (WHO) estimates to have sensitivity of 66% and specificity of 87%. For women living with HIV, the AUC of AVE was 0.91 (95% CI = 0.88 to 0.93), and among those testing positive for high-risk HPV types, the AUC was 0.87 (95% CI = 0.83 to 0.91).

**Conclusions:** These results demonstrate the feasibility of utilizing AVE on images captured using a commonly available smartphone by screening nurses and support our transition to clinical evaluation of AVE’s sensitivity, specificity, feasibility, and acceptability across a broader range of settings. The performance of the algorithm as reported may be inflated, as biopsies were obtained only from study participants with visible aceto-white cervical lesions, which can lead to verification bias; and the images and data sets used for testing of the model, although “unseen” by the algorithm during training, were acquired from the same set of patients and devices, limiting the study to that of an internal validation of the AVE algorithm.

## II. Background

Each year, cervical cancer kills more than 300,000 women, with most deaths occurring in low- and middle-income countries (LMIC) where the prevention and therapeutic infrastructure is extremely limited.^1,2^ When screenings are conducted, Visual Inspection with Acetic Acid (VIA) is the most commonly used screening tool for the detection of precancerous lesions; however, it is a relatively inaccurate method with significant heterogeneity of performance across providers, with sensitivity ranging from 22% to 91% and specificity ranging from 47% to 99% in published studies. ^3^ Widespread use of VIA is largely due to its low cost and the perceived simplicity with which it can be administered, even in rural areas. It also facilitates same-visit treatment of precancerous lesions. For these reasons, it has been integrated into many national cervical cancer screening efforts.

The 2021 World Health Organization (WHO) cervical cancer screening and treatment guideline recommends the use of human papillomavirus (HPV) DNA testing as a primary screening method where feasible. With a negative predictive value close to 100%,^4^ a negative HPV DNA test result offers considerable assurance that a woman will not soon develop precancerous lesions, lengthening the intervals for when women need to return for re-screening. Women can also collect their own samples through a vaginal swab, avoiding the need for a pelvic exam with a speculum. However, HPV tests are not currently affordable for widespread use in most low-income countries, and practical point-of-care HPV testing solutions are not yet available, posing major barriers to scale-up and sustainability.

VIA can be used by providers to determine whether women who test HPV positive from both the general population and those living with HIV should be treated and/or what type of treatment is appropriate. When using the “screen and treat” approach, WHO recommends treating all women who test HPV positive. In such instances VIA can be used to determine the most appropriate treatment, depending on the characteristics of the cervical transformation zone and cervical precancerous lesion, if the latter is visible. When using the “screen, triage, and treat” approach, WHO recommends triaging all women who test HPV positive with a visual test, such as VIA, to determine if treatment is mandated and what type is appropriate.

For these reasons, visual evaluation will remain a fundamental component of national screening and treatment programs in LMICs for the foreseeable future, especially as local governments and their global public health partners work to scale up secondary prevention services to accelerate the elimination of cervical cancer as a public health problem. Still, for visual evaluation to achieve its intended impact at sustained scale, innovation is needed to improve accuracy and reliability.

A proposed screening method termed Automated Visual Evaluation (AVE), which uses a machine learning algorithm to analyze an image of the cervix and provides a result indicating whether precancerous lesions are likely present or absent, has the potential to fill this gap. To use AVE, a provider takes a picture of the cervix using a standard smartphone, runs the algorithm on the mobile phone processor, and then receives a ‘positive’ or ‘negative’ result in less than one minute. AVE is intended as a decision-making aid to assist providers conducting VIA.

Results published in 2019 of an AVE algorithm trained and tested on data from a longitudinal study in Costa Rica demonstrated the proof of principle for AVE^5^ with an area under the curve (AUC) of 0.91, which translated into an estimated sensitivity of 97.7% and specificity of 84% in identifying precancerous lesions and cancer (cervical intraepithelial neoplasia grade 2 or greater; “CIN2+”) among women 25-49 years old. The accuracy of the AVE algorithm proved superior to clinical opinion or conventional cytology; however, generalizability of the Guanacaste study is limited by the relative homogeneity of the dataset. All women tested were from Costa Rica, which has a significantly lower HIV prevalence than other LMICs, particularly in Africa. Further, the Guanacaste analysis was performed using high-quality images of standard angle, light, and focus, captured by a small team of highly trained nurses using a cerviscope—a fixed-focus, ring-lit film camera that has since been discontinued. To be used at scale, an AVE algorithm would need to work on a more manageable and affordable device, and on images captured by health workers with a range of experience and training levels.

This article describes the results of our effort to adapt AVE for use on a commercially available and affordable smartphone, with the machine learning training, validation, and testing of AVE using smartphone images collected by providers from women in Zambia. An approach combining AVE and HPV genotyping, based on a subset of the data from this same study, was previously published^6^.

## III. Materials and Methods

### Clinical Methods

Our study was nested within Zambia’s public sector cervical cancer prevention service platform. More than 8,000 women aged 25-55 were prospectively recruited from among the population of women attending one of eight Ministry of Health public health facilities in Lusaka for routine care from October 2019 through December 2021. Zambia was chosen as the site for this study based on the presence of sophisticated clinical and patient management infrastructure—the product of a fifteen-year investment by the Zambia Ministry of Health (MOH), local partners, and external donors, including the US President’s Emergency Plan for AIDS Relief (PEPFAR).

In 2005, the VIA screening method was launched in three government-operated primary healthcare clinics co-housed with HIV/AIDS programs funded by PEPFAR, in the capital city of Lusaka. As health promotion activities expanded and community health workers addressed local myths and misconceptions about cervical cancer, the demand for screening services increased, creating opportunities for cancer prevention as well as clinical infrastructure and workforce challenges.

To improve service efficiency, a local wireless telecommunications platform was developed and implemented. Using ‘digital cervicography’ (DC) as an adjunct to VIA, providers (mostly nurses and nurse midwives) were trained to routinely capture images of the cervix with a commercial-brand digital camera outfitted with a macro-lens. The images were magnified on a television monitor for detailed examination by providers and for patient education. Referred to as ‘electronic cervical cancer control’ (eC3), the novel solution facilitated immediate distant consultation with local experts, monitoring and evaluation of clinical decisions made by providers, digital image-pathology correlations, and medical records documentation. Subsequent research has shown the eC3 DC approach helped improve sensitivity for precancerous cervical lesions among women living with HIV (WLHIV) compared to naked-eye VIA.^7^ Physician workforce shortages were ameliorated by task-shifting procedures (cryotherapy, thermal ablation, loop electrosurgical excision procedure (LEEP)/large loop excision of the transformation zone (LLETZ), punch biopsy) from gynecologists to nurses and clinical officers.^8^

With strong high-level advocacy from the Office of the First Lady of Zambia, leadership from the Ministry of Health, a robust community education program, and an innovative collaboration of traditional leaders (e.g., local tribal chiefs, traditional healers, traditional marriage counselors), a matrix of nurse-led same-day ‘screen-treat’ services (VIA, thermal ablation, loop electrosurgical excision procedure (LEEP)/large loop excision of the transformation zone (LLETZ), punch biopsy) was successfully scaled to 160 public health clinics across all of the country’s 10 provinces. Altogether, these investments in human resource development, alongside electronic data collection and public sector pathology laboratory infrastructure, have enabled Zambia’s cervical cancer prevention platform to now support extramurally funded clinical research, including randomized clinical trials and our scientific investigations involving AVE algorithms.

The study was designed to collect at least 750 histopathology-confirmed patient images of precancer, as this was judged to be a sufficient target for machine learning training and validation experiments. Because precancerous lesions are relatively rare, nesting the study in nurse-led clinics in which same-day “screen-treat” services included electrical excision procedures (LEEP/LLETZ) allowed us to meet the target number of CIN2+ images. In the study, more than 8,000 women were prospectively recruited from among a population of women receiving routine care from one of eight public health facilities, seven in Lusaka and one in Kitwe, during the period of October 2019 through December 2021. The women were eligible to participate if they were aged 25-55 years (age range recommended for cervical cancer screening by the MOH) and able to understand and willing to sign a written informed consent form. Women were excluded if they were pregnant or had a history of hysterectomy, trachelectomy, or treatment of cervical precancer or cancer. Women were enrolled consecutively at each site.

After eligibility and consent were confirmed, study participants were offered HIV and HPV testing. Following HPV sample collection using the COPAN FLOQSwab, cervical cancer screening was performed using ‘digital cervicography’ (DC) as an adjunct to VIA in accordance with guidelines established for the government-operated ‘screen and treat’ clinics. For DC-VIA, the cervix was lavageed with a cotton swab soaked in 5% acetic acid by nurse-providers. After a waiting period of 2-3 minutes, an image of the cervix was obtained using a digital camera, then magnified to (1) identify the transformation zone (TZ) type, (2) determine the presence and characteristics of aceto-white lesions, and (3) decide the appropriate form of management. The digital camera cervical image was then shown to and discussed with the participant along with the recommended management plan.

For this study, with a goal to train the AVE algorithm only and not for informing clinical care decisions, additional images of the cervix (typically three images per participant) were obtained with a smartphone in quick succession after the digital camera image was captured. Two smartphone models, the Samsung Galaxy J8 and Samsung Galaxy A21s, readily accessible and affordable in Zambia at the time of data collection, were used to collect these images. A commercially available smartphone app-Box.com - was used to capture and store the images on the smartphones. Typically, only one model of smartphone was used for each participant, but some participants had images captured by providers using both models during the transition period while switching phone models. When lesions were identified, participants were treated in line with the Zambia MOH cervical cancer screening guidelines. Specifically:

1. Participants whose transformation zones (TZ) and any aceto-white lesions, if present, were purely ectocervical (TZ type l), and occupied less than three-fourth of the transformation zone and were without features of invasive cancer, were offered ablative therapy using thermal ablation, performed using the Liger handheld cordless coagulator. ^9^
2. Participants whose transformation zones extended into the endocervical canal (TZ typell/lll) or whose lesions occupied more than three-fourth of the transformation zone, were offered LEEP/LLETZ. ^10^
3. For participants whose lesions were suspicious for cancer, a punch biopsy was performed.

As an additional study procedure, a punch biopsy was performed prior to ablation on all women with aceto-white lesions and eligible for ablative therapy.

For our study reference, we leveraged cervical histopathology to confirm the presence of cervical precancerous lesions. Biopsy samples were taken from all women who screened positive on digital cervicography (DC) or VIA. Histopathologic analysis was performed on all tissue (punch biopsy and LEEP/LLETZ)) specimens, with results reported as normal, benign lesion, CIN1, CIN2, CIN3, micro-invasive cancer, or invasive cancer. Study participants found to have invasive cancer on histopathology were referred to the Gynecologic Oncology Unit of University Teaching Hospitals (UTHs) - Women and Newborn Hospital in Lusaka for appropriate staging and treatment. For all study participants, screening (and treatment, if applicable) was followed up with routine screening services in the same government-operated clinics, per the routine standard of care.

HPV testing was performed with the Beckton Dickinson (BD) Viper LT system, using the BD Onclarity™ HPV Assay as per manufacturer’s specifications and standardized performance instructions. The BD Onclarity™ HPV Assay is an automated laboratory test that detects DNA (deoxyribonucleic acid) from 14 high risk HPV types that are associated with cervical cancer. The test specifically identifies HPV types 16, 18, 31,45, 51, 52 while concurrently detecting types 33/58, 35/39/68, and 56/59/66. For the purpose of this analysis, the results were classified as being high-risk positive or negative. The assay was performed at the Women’s and Newborn Hospital/University of North Carolina HPV Laboratory located on the campus of UTH.

The study protocol was reviewed and approved by the University of Zambia Biomedical Research Ethics Committee, Zambian National Health Research Agency, and the University of North Carolina Institutional Review Board, in addition to scientific and programmatic reviews by various committees at funding agencies. All study participants provided written informed consent. Women who declined or were unable to provide written informed consent were offered screening and treatment for precancerous lesions in line with the standard of care in Zambia. This study was designed to collect data for training an AVE algorithm as well as evaluating the performance of the algorithm; as diagnostic accuracy is evaluated, this study is being reported in accordance with the Standards for Reporting Diagnostic accuracy studies (STARD) checklist (STARD supplement checklist) ^11^.

### Machine Learning Methods

Machine learning leverages the abilities of computers to analyze large datasets and draw conclusions from patterns. To train an AVE algorithm to recognize cervical precancer cases accurately, thousands of images representing control status or histopathology-confirmed case status were presented to the algorithm. To diagnose cervical precancer and cancer, histopathology is considered the gold standard test. The inputs to train our AVE model included the set of cervical images and the ground truth label for each image. The ground truth label was a binary input - case vs. control – with case representing CIN2, CIN3, or cancer (‘CIN2+’) and control representing images that had a histopathology result less severe than CIN2 (‘< CIN2’) or images that were negative on DC-VIA (and therefore represent participants who did not receive a biopsy).

Smartphone images were labeled with the study ID (and no other identifiers) and uploaded directly from the smartphones to our study’s machine learning team via the Box.com App. Linked, de-identified participant data with key demographic and clinical details (age, HIV status, DC-VIA result, HPV test result, histopathology result if applicable) were shared electronically with the machine learning team via Excel files.

Our study objective was to train, validate, and test an algorithm with high discrimination between cases and controls. This paper details the AVE algorithm developed and tested using images from the Samsung A21s smartphone model. As detailed in Figure 1, participants were excluded if they (1) did not have a good quality A21s smartphone image or (2) had screened positive but did not have a histology result. This resulted in 13484 A21s images, representing 5188 participants. These A21s images comprise the data set used for the training and evaluation of AVE algorithm reported here. The Samsung J8 images were used in preliminary development as described later in this section and in the supplementary.

**Figure 1.**
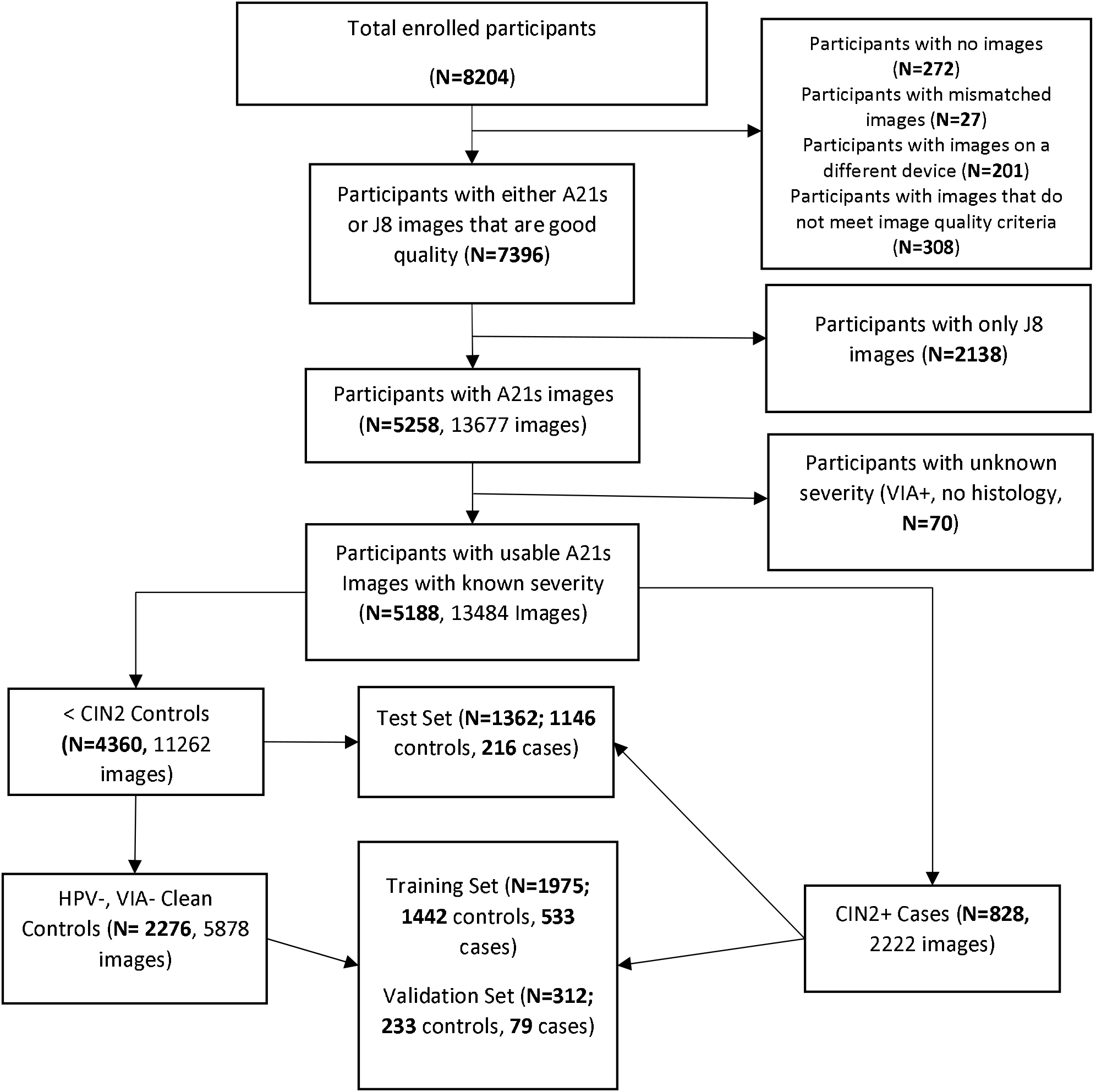
Participants and training/test splits.

**Figure 2.**
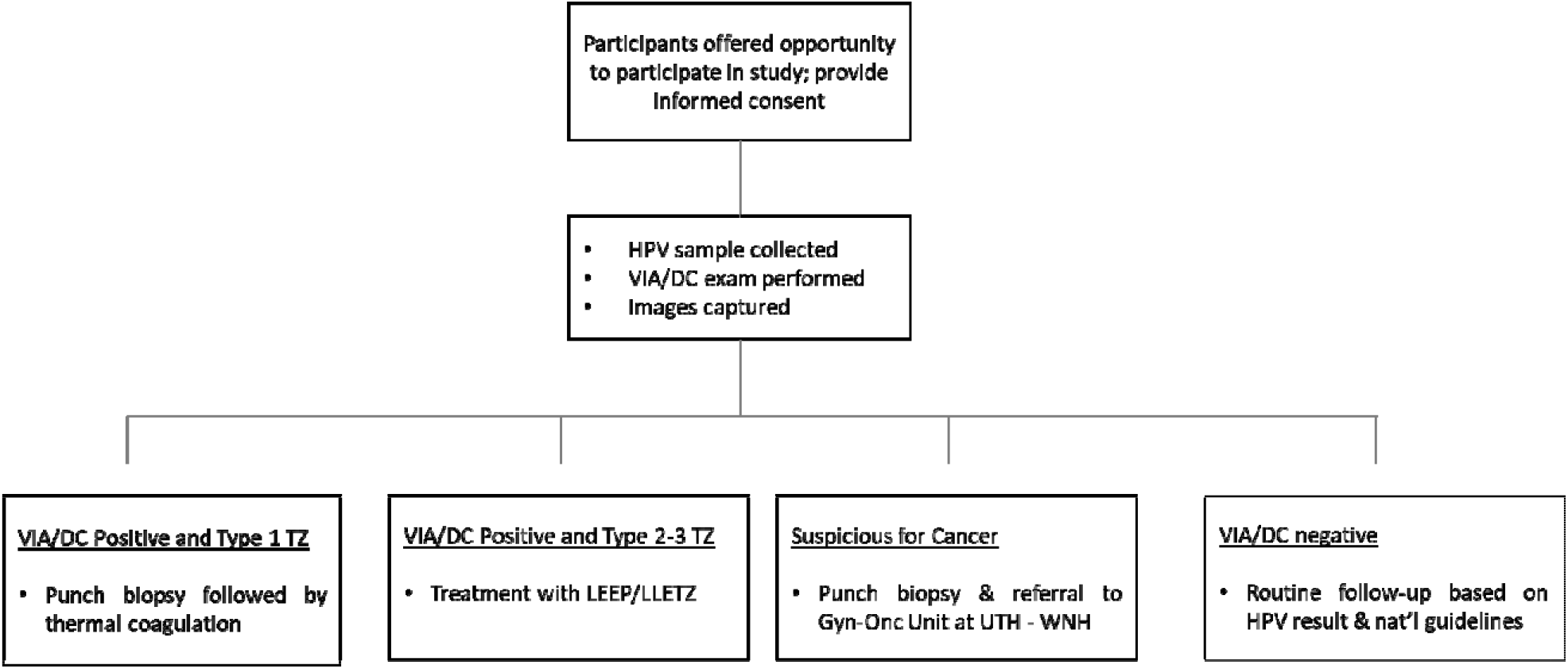
Participant workflow.

In line with standard machine learning procedures, images were divided into a training set (65% of images), validation set (10% of images) and test set (25% of images) randomly. This division was done at the participant level, such that if a participant had more than one image in the database all such images were put into the same set (training, validation, or test) to prevent data leakage.

The machine learning methods used in this study match the methods previously reported in Hu et al (2020)^12^, which showed that the AVE cervical precancerous lesion detection algorithm can be refactored to run on the limited computing power of a smartphone with minimal impact on performance using the RetinaNet framework^13^. A key difference between the two studies is that the work presented here is based on smartphone images whereas the Hu *et al* work was based on digitized cervigram images. Other differences include our study rescaling the images to a width of 900 pixels and height of 1200 pixels (with most images maintaining the original aspect ratio) and selecting a learning rate of 2e-5.

Another significant difference involved our experimenting with different subsets of cases and controls used for training the AVE algorithm. Each image is linked to the corresponding HPV test result, VIA result, and (where applicable) histology result. Based on these test results, participants can be divided into several distinct categories. For our study, participants were divided into the categories shown in Table 1. For example, a study participant with an HPV negative and VIA negative result was labeled as “Category 1a” and a study participant with histology CIN3 was labeled as “Category 3b”. While the overall target of our AVE model is to separate all CIN2+ subjects from all <CIN2 subjects, this finer categorization allows training models using specific categories, as described below. It also allowed us to estimate performance in different settings such as general population screening or HPV triage. It is possible that this finer categorization might enable training multiclass (rather than binary) models, and even finer subcategorization, for example through stratification of study participants by HPV genotype, is possible, but our current study did not explore these.

**Table 1.** Participant categorizations.

|  |  |  |
| --- | --- | --- |
| 1 | Normal |  |
|  | 1a | [HPV-, VIA-] |
|  | 1b | [HPV-, VIA+, Histology normal] |
| 2 | Low risk |  |
|  | 2a | Low risk due to HPV only |
|  |  | 2a.1 [HPV+, VIA-] |
|  |  | 2a.2 [HPV+, VIA+, Histology normal] |
|  | 2b | Low risk due to Histology only: [HPV-, CIN1] |
|  | 2c | Low risk due to HPV and Histology: [HPV+, CIN1] |
| 3 | Precancer |  |
|  | 3a | CIN2 |
|  |  | 3a.1 CIN2, HPV+ |
|  |  | 3a.2 CIN2, HPV- |
|  | 3b | CIN3 |
|  |  | 3b.1 CIN3, HPV+ |
|  |  | 3b.2 CIN3, HPV- |
| 4 | Cancer |  |

During initial model development, we trained binary models on the Samsung J8 images splitting the controls and cases in several different ways, as described in Supplement S1. Each model used a different definition of case versus control during the training and validation process. Importantly, while not all categories of images were used in the training and validation sets, all categories were used in the test set to assess each model’s ability to distinguish CIN2+ from <CIN2 across a full range of potential disease states. The categorization split producing the model that performed best (defined as the highest AUC) on a Samsung J8 test set was that in which training controls were limited to HPV- and DC-VIA-images (category 1a) and the training cases included all CIN2+ images (categories 3 and 4). This same split across categories was then used to train models with Samsung A21s images.

To explore device portability of the AVE models (how well a model trained on data from one device performs on data from another device), we evaluated the performance of models trained with J8 data only, A21s data only, and combinations of varying amounts of J8 and A21s data on the test sets from each smartphone. This work is described in detail in Supplement S2.

### Analysis

We analyzed results on Samsung A21s images to support subsequent AVE validation studies on that device.

The primary measure of diagnostic accuracy is the Receiver Operating Characteristic (ROC) curve and its summary statistic, area under the curve (AUC).

AVE’s **sensitivity** is defined as:

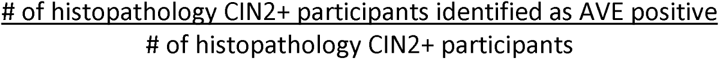

AVE’s **specificity** is defined as:

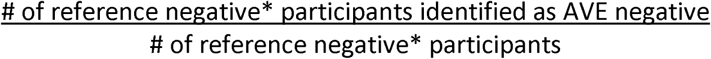

*\*Reference negative is defined as EITHER histopathology result of no precancer/cancer (i.e*., *<CIN2) OR DC-VIA-negative screening result. This consists of the entirety of categories 1 and 2 from Table 1*.

Study participants with a DC-VIA-positive result, but lacking a histopathology result, were not included in the AVE test set and performance analysis. Participants without an HPV result or VIA result were included in the AVE test set and performance analysis when a histopathology result was available.

AVE is intended as a primary screening tool or as a triage test for women who are HPV-positive. Our training and testing data from Zambia were enriched with data from patients that underwent LEEP/LLETZ based on an initial screen. This was designed to help our study team collect a sufficient absolute number of images of CIN2+ to train the AVE algorithm. However, it also resulted in an image set with higher severity relative to a typical screening setting. In response, we “rebalanced” our test set to enable an accurate measure of AVE’s performance in a primary screening or HPV+ triage setting that would be most typical for the geographies where AVE is intended to be used. See Supplement S3 for a description of this rebalancing process.

We also analyzed the results of AVE to validate different use cases; specifically, the use of AVE as a triage test for women who are HPV positive and the use of AVE among WLHIV. Tables 2 and 3 show the number of participants and images in the test sets for each of these use cases.

**Table 2.** Participants and image count of the AVE test set by HPV status.

| HPV status | # of participants | # of images |
| --- | --- | --- |
| HPV negative | 715 | 1825 |
| HPV positive | 516 | 1292 |
| HPV unknown | 131 | 344 |

**Table 3.** Participants and image count of the AVE test set by HIV Status.

| **HIV status** | **# of participants** | **# of images** |
| --- | --- | --- |
| HIV negative | 607 | 1480 |
| HIV positive | 733 | 1932 |
| HIV unknown | 22 | 49 |

### Public and patient involvement

Patients (i.e., participants of the study) were not involved in the design of this phase of the research study. However, we anticipate client involvement as we conduct implementation research because provider, client, and community partner acceptance will be key to the scale up and sustained use of AVE in LMICs.

## IV. Results

### Clinical results

Of the 8,204 women aged 25-55 who were screened with HPV testing and VIA with digital cervicography (DC-VIA), 4,390 (54%) were HIV positive; 3,388 (44%) were positive for HPV; and 3,149 (38%) were positive on DC-VIA. Of the participants who were DC-VIA-positive, 1,024 (33%) women were treated with thermal ablation and 1,807 (57%) were treated with excision. (A further 318 women (10%) were suspicious for cancer at the time of screening and referred to Gynecologic Oncology Unit at UTH-Women and Newborn Hospital for further evaluation and cancer treatment, as required.) Of the 3,121 women who underwent histopathology analysis, 817 (26%) were negative; 566 (18%) had CIN1; 1,337 (43%) had CIN2/3; 345 (11%) were found to have invasive cervical cancer; and 56 (2%) had unsatisfactory or invalid histopathology results.

### Machine learning results

We evaluated the accuracy of the AVE algorithms trained and tested using Samsung J8 images with various categorization splits (as described in the methods section and Supplement S1), for the detection of CIN2+ using Area Under the Receiver Operating Characteristic Curve (AUC). The AUCs of the relevant J8 algorithms are shown in Supplement S1 and based on this, the final model utilizing A21s images was trained on a split with A21s case images consisting of all CIN2+ categories, but A21s control images were limited to participants testing negative on DC-VIA and HPV test.

At the image level, the AUC of the model trained on A21s images when tested on the A21s test set was 0.91 (95% CI = 0.90 to 0.92). To get participant-level predictions, we chose the image with a predicted confidence score closest to zero (if the image was detected as normal cervix) or one (if the image was detected as precancerous cervix) for each participant. In other words, we chose the image with the “maximum confidence”. We applied this method to the test set predictions, achieving a participant-level AUC of 0.91 (95% CI = 0.89 to 0.92). This method represents a scenario in which up to three images are captured from each subjects and used in evaluation. We also explored the consistency of the output for each image. For participants in the test set with at least two suitable images, there was agreement of the precancer classification for all images for 88% of participants.

We then adapted the test set results to the general population setting, described in more detail in supplement S3. This resulted in a participant-level AUC of 0.91 (95% CI = 0.89 to 0.93), with the threshold based on maximizing Youden’s index resulting in sensitivity of 85% (95% CI = 81% to 90%) and specificity of 86% (95% CI = 84% to 88%). We also evaluated performance among women testing positive for high-risk HPV, achieving an AUC of 0.87 (95% CI = 0.83 to 0.91), and among WLHIV, achieving an AUC of 0.91 (95% CI = 0.88 to 0.93). The ROC curves for general population screening, HPV triage, and among WLHIV settings are shown in Figure 3.

**Figure 3.**
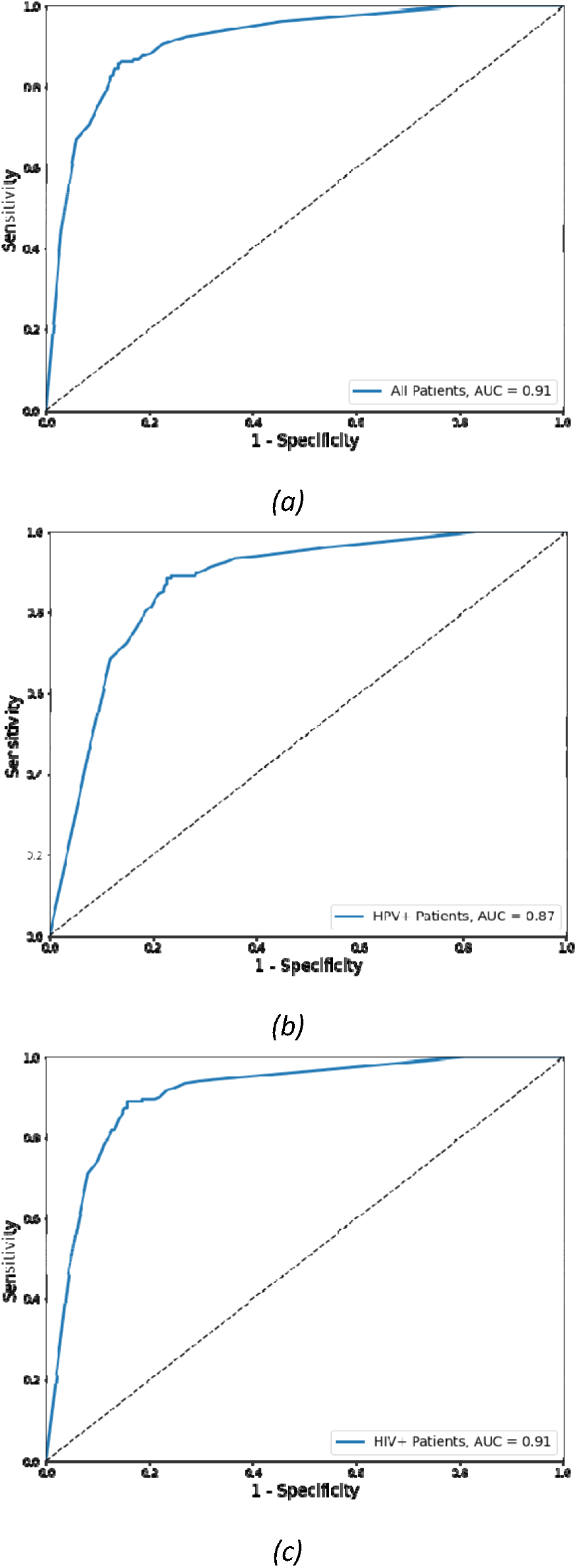
ROC curves of A21s AVE model for participant level of (a) general population screening AUC = 0.91; (b) HPV+ triage population screening AUC = 0.87 and (c) population screening for WLHIV AUC = 0.91.

The results of the device portability experiments are presented in Supplement S2. Briefly, the model trained with only A21s data that produces an AUC of 0.91 on the A21s test set produces an AUC of 0.79 on a J8 test set, and a model trained with only J8 data produces an AUC of 0.83 on the A21s test set. Adding training data for the device under test (“primary device”) improves the AUC, as expected, and less data from the primary device is needed if accompanied by data from a secondary device than if limited to only data from the primary device. For example, for the A21s test set, reaching an AUC of 0.87 when training with only A21s data was not achieved until training with 212 cases and 578 controls (at the patient level). However, when combined with a large J8 training data set (432 cases, 609 controls), the same AUC is achieved by adding only 53 A21s cases and 144 A21s controls.

## V. Discussion

In this work, we developed AVE, an AI-based algorithm that can be adapted for running on a smartphone that can detect CIN2+ precancerous lesions on images of the cervix captured with low cost, commonly available smartphones. The study demonstrated that the performance of AVE for primary cervical cancer screening was high as a primary screening method for all women (AUC = 0.91, 95% CI = 0.89 to 0.93) and WLHIV (AUC = 0.91, 95% CI = 0.88 to 0.93) and was also high when used as a triage when women screening positive for high-risk HPV (AUC = 0.87, 95% CI = 0.83 to 0.91). These results underscore AVE’s potential as a highly accurate, accessible tool for screening precancerous cervical lesions and the technology’s broader potential to help accelerate the *Global Strategy to Eliminate Cervical Cancer as a Public Health Problem*.^14^

AVE-assisted VIA holds promise as an effective primary screening tool within geographies where HPV testing is not yet available to all women in need of screening. With a sensitivity of 85% (CI=81% to 90%) and specificity of 86% (CI = 84% to 88%) at a threshold that maximizes the Youden’s index, the AVE algorithm represents a considerable improvement in sensitivity over VIA alone that has been reported to have a sensitivity of 66% (95% CI: 61%-71%) in WHO’s meta-analysis, with “extreme heterogeneity” in reported results across studies (22% to 91%).^3^ If the use is further clinically validated, the use of AVE-assisted VIA as a primary screening test may enable women to be screened and treated for precancerous lesions by a more reliable and objective test in a single visit, a critical element of woman-centered care that increases the likelihood that women in need of treatment can access it. AVE-assisted VIA can also be used as a triage test for women who are HPV-positive in countries that use a ‘screen-triage-treat’ approach with HPV screening as the primary test.

This study, focused on data collected from experienced clinicians in Zambia, was an important first step to develop the algorithm from a clinical environment representing the proposed use case. It is important to highlight that the results presented are from internal validation, i.e. the reported results are based on testing the algorithm on a population drawn from the same study as the training data. External validation of the AVE algorithm, i.e. testing on data from a different study populations and settings, is needed to fully understand the generalization performance of the algorithm. Prior work has shown that performance typically drops in external validation.^15^ While we do not have data to suggest that cervical appearance varies based on women’s ethnicity, race, or geography of origin, previous research has shown that the prevalence of underlying conditions with the potential to affect cervical appearance – such as female genital schistosomiasis and chronic cervicitis – can vary by setting. Clinic environments, such as variations in lighting, exam room set-up, quality of acetic acid, and provider skill, may also vary by setting and could potentially affect algorithm performance.

An additional limitation is the likelihood of verification bias since histopathological results are only available from women who screened positive by DC-VIA, thus all women who screened negative on DC-VIA are assumed to be negative for CIN2+ for this work. Although VIA with DC has shown higher sensitivity than VIA^7^, the possibility of incomplete disease ascertainment cannot be eliminated. This limitation means the sensitivity estimates reported here are likely inflated ^16^. In a follow-up study in which all HPV-positive women are referred to histopathology (unpublished, analysis is ongoing) there is evidence of decreased AVE sensitivity due to VIA-negative, histopathology-confirmed precancers..

Future versions of AVE with additional optimizations are also under development. This work presents the results of an object detection model that is configured to produce a binary output: positive or negative for cervical precancer/cancer. Other algorithm approaches (such as multiclass, Bayesian, or evidential models) exist and may be beneficial, potentially combined with an additional test like HPV testing, to better deal with uncertain results and stratify individuals by risk level.^6,15^ In addition, it may be feasible, for example, to develop an additional algorithm that can determine eligibility for ablative vs. excisional treatment, based on the anatomy of the cervical transformation zone and the position and size of any lesions.

Device portability of AVE algorithms is still an unsolved problem as both our results and prior work^6,15^ suggest that training data from a new device is needed to optimize performance on that device. This is particularly challenging for smartphones which have relatively short lifespans. Ongoing efforts are underway to explore the feasibility of porting an algorithm to a new smartphone and whether the required number of training images per device decreases as additional device types are included in training. Alternatively, a dedicated medical device with fixed imaging hardware and longer production cycle could help ameliorate this problem.

While this work continues at pace, AVE holds enormous potential for improving the accuracy and consistency of visual inspection for precancerous cervical lesions. It could help shape a paradigm shift in the effort to screen and treat women before cervical cancer develops, particularly in LMICs.

## VI. Other information

The study was supported by Unitaid, Global Health Labs, Inc (www.ghlabs.org) and the US National Cancer Institute (NCI) (grant UH3CA202721). We would like to acknowledge the support of the current or former colleagues of Global Health Labs, Inc: Sheana Creighton, Shannon Kuyper, Ben Wilson, and Courosh Mehanian, and current or former colleagues in Zambia involved with the study management including Namakau Nyambe, Bridget Lumbwe, Emmanuel Muzumbwe, and Chusi Sikanyika.

Opinions expressed by the authors are their own and this material should not be interpreted as representing the official viewpoint of the U.S. Department of Health and Human Services, the National Institutes of Health, or the National Cancer Institute.

The lead author affirms that the manuscript is an honest, accurate, and transparent account of the study being reported; that no important aspects of the study have been omitted; and that any discrepancies from the study as originally planned have been explained.

## VII. Data availability statement

The data presented in this study are available upon reasonable request to the authors.

