## Supplements for "Internal Validation of Automated Visual Evaluation (AVE) on Smartphone Images for Cervical Cancer Screening in a Prospective Study in Zambia"

### Supplement S1. Initial experiments based on Samsung J8 images.

As described in the main text, initial AVE models based on J8 images were trained using different severity subcategories of participants as cases and controls. Participant splits of the initial experiments are shown in Table S1.1, along with a brief description of the training set and/or what the model was trained to distinguish.

Table S1.1. Categorization splits for initial J8 AVE algorithm models trained.

| Categorization split # | Image category comparisons^*^ | Model explanation |
| --- | --- | --- |
| 1 | [1] vs. [3,4] | Model was shown only normal control images during training; low risk were not used. |
| 2 | [1,2] vs. [3,4] | Model was shown both normal and low risk control images during training. |
| 3 | [1] vs. [2,3,4] | Tested feasibility of distinguishing normal from all other image types (including low risk). |
| 4 | [1a] vs. [3,4] | Model was only shown “clean control” (HPV-/VIA-) images during training. |
| 5 | [1,2a] vs. [3,4] | Model was shown normal and low risk <CIN1 as controls during training; CIN1 was not used. |
| 6 | [1a,2a1] vs. [3,4] | Model was shown only VIA- controls during training. |
| 7 | [1b,2b,2c] vs. [3,4] | Model was shown only VIA+ or CIN1 images during training to test how a model could distinguish VIA+ controls from CIN2+ cases. |

^*^See main text, Table 1 for participant category definitions.

The seven models in Table S1.1 targeted the main AVE goal of distinguishing CIN2+ from <CIN2 participants. The resulting AUCs of these models, trained and tested only with J8 images, are shown in Table S1.2. Split 4 was trained and validated with women who were HPV-/VIA- (as the “control”) and CIN2+ (as the “case”), which had the highest AUC when tested against a holdback set of test images of all severity categories. We observed that training with just clean controls (split 4) produced a strong AUC of 0.88 on the broader test set. Adding more control groups never improved the AUC. In fact, it decreased sensitivity and specificity, so we chose split 4 for the A21s experiments described in the main text.

Table S1.2. J8 AVE algorithm test results of models trained with different control/case categorization splits.

| Categorization split # | J8 test set AUC (non CIN2+ vs. CIN2+, including all the severity groups) |
| --- | --- |
| 1 | 0.87 |
| 2 | 0.86 |
| 3 | 0.83 |
| 4 | 0.88 |
| 5 | 0.88 |
| 6 | 0.87 |
| 7 | 0.80 |

### Supplement S2. Algorithm portability between smartphone models

While the model described in the main text was trained with only Samsung A21s data, further work to test device portability was conducted with both Samsung A21s and Samsung J8 data. The A21s training, validation, and test data used is the same as shown in Figure 1 of the main text, while the J8 data was similarly split at a 65% training : 10% validation: 25% test ratio, ensuring that any participants who had images from both phones were in the same split (Table S2.1). For instance, any J8 images from a participant with A21s validation images was put in the J8 validation set. Also consistent with the main text, only HPV-, VIA- clean controls were included in the J8 training and validation sets.

*Table S2.1*: Training and validation participant counts for the two phone models

|  | Training participants (# cases, # clean controls) | Validation participants (# cases, # clean controls) |
| --- | --- | --- |
| Samsung A21s | 1975 (533, 1442) | 312 (79, 233) |
| Samsung J8 | 1041 (432, 609) | 147 (68, 79) |

*Table S2.2*: Number of participants per phone in the training and validation set at each percentage level used in the portability experiments.

|  | **Samsung A21s participants** | | | | **Samsung J8 participants** | | | |
| --- | --- | --- | --- | --- | --- | --- | --- | --- |
| **% of data utilized** | **Training # CIN2+** | **Training # Clean Controls** | **Validation # CIN2+** | **Validation # Clean Controls** | **Training # CIN2+** | **Training # Clean Controls** | **Validation # CIN2+** | **Validation # Clean Controls** |
| **10%** | 53 | 144 | 8 | 23 | 43 | 61 | 7 | 8 |
| **20%** | 106 | 288 | 16 | 46 | 86 | 122 | 14 | 16 |
| **40%** | 212 | 576 | 32 | 92 | 172 | 244 | 28 | 32 |
| **60%** | 319 | 864 | 47 | 139 | 258 | 366 | 42 | 48 |
| **80%** | 426 | 1153 | 63 | 186 | 345 | 488 | 56 | 64 |
| **100%** | 533 | 1442 | 79 | 233 | 432 | 609 | 68 | 79 |

In addition to testing how well a model trained on data from one device would perform on data from the 2nd device, we aimed to explore how the model performance changed as increasing amounts of data from the 2^nd^ device was added. Each phone’s training and validation sets were randomly split into ten nearly equal parts, known as folds, at the participant level. This division was done while ensuring a balanced representation of different severity levels - clean controls, CIN2, CIN3, and cancer - in each fold. Using these folds, we incrementally built datasets in 10% steps. For instance, utilizing only the first fold equates to using 10% of the data, the first two folds represent 20%, and this pattern continues until the entire dataset is included. This method facilitated the exploration of various data percentage combinations from each phone, while maintaining consistency in the data used across all portability experiments.

We trained models utilizing every combination of 0%, 10%, 20%, 40%, 60%, 80%, and 100% of the participants’ data for each phone. The numbers of participants included in the training and validation sets for each experiment are recorded in Table S2.2. Each model was then run on both the A21s and J8 test sets, in which, as in the main text, negatives included all participants that were not classified as CIN2+ (i.e. not limited to only clean controls). ROC AUC values for the A21s test set are in Table S2.3, and for the J8 test set in Table S2.4.

*Table S2.3:* Comparison of ROC AUC Values for Models Trained with Varied Proportions of A21s and J8 Participants on A21s test set

|  |  | **% of A21s training and validation participants** | | | | | | |
| --- | --- | --- | --- | --- | --- | --- | --- | --- |
|  |  | **0%** | **10%** | **20%** | **40%** | **60%** | **80%** | **100%** |
| **% of J8 training and validation participants** | **0%** |  | 0.59 | 0.85 | 0.87 | 0.90 | 0.90 | 0.90 |
|  | **10%** | 0.61 | 0.82 | 0.86 | 0.88 | 0.90 | 0.89 | 0.90 |
|  | **20%** | 0.64 | 0.84 | 0.86 | 0.87 | 0.90 | 0.90 | 0.90 |
|  | **40%** | 0.74 | 0.85 | 0.87 | 0.89 | 0.90 | 0.88 | 0.91 |
|  | **60%** | 0.86 | 0.87 | 0.87 | 0.89 | 0.91 | 0.89 | 0.90 |
|  | **80%** | 0.82 | 0.85 | 0.87 | 0.89 | 0.89 | 0.90 | 0.90 |
|  | **100%** | 0.83 | 0.87 | 0.88 | 0.89 | 0.88 | 0.89 | 0.89 |

*Table S2.4:* Comparison of ROC AUC Values for Models Trained with Varied Proportions of A21s and J8 on J8 test set

|  |  | **% of A21s training and validation participants** | | | | | | |
| --- | --- | --- | --- | --- | --- | --- | --- | --- |
|  |  | **0%** | **10%** | **20%** | **40%** | **60%** | **80%** | **100%** |
| **% of J8 training and validation participants** | **0%** |  | 0.52 | 0.75 | 0.80 | 0.81 | 0.82 | 0.79 |
|  | **10%** | 0.44 | 0.82 | 0.84 | 0.87 | 0.87 | 0.87 | 0.83 |
|  | **20%** | 0.79 | 0.85 | 0.88 | 0.86 | 0.86 | 0.88 | 0.86 |
|  | **40%** | 0.82 | 0.84 | 0.86 | 0.87 | 0.85 | 0.83 | 0.87 |
|  | **60%** | 0.86 | 0.85 | 0.86 | 0.86 | 0.88 | 0.85 | 0.86 |
|  | **80%** | 0.87 | 0.85 | 0.85 | 0.86 | 0.87 | 0.88 | 0.88 |
|  | **100%** | 0.87 | 0.87 | 0.87 | 0.88 | 0.87 | 0.87 | 0.87 |

As can be seen in Table S2.3, a model trained with only J8 data shows reasonable portability to the A21s test set, with an AUC of 0.83. However, adding even a small quantity (10%) of A21s data boosts the performance to 0.87, and adding the full A21s data to 0.89. In contrast, in the absence of any J8 data, 40% of the A21s data is needed to achieve an AUC of 0.87. Similarly, on the J8 test set, the AUC trends from 0.79 with only A21s training data to 0.86 with just 20% of the J8 training data, whereas achieving this AUC in the absence of A21s data requires 60% of the J8 data.

Another interesting finding is that near-maximal performance on their respective test sets can be achieved with just 60% of A21s data (0.90 AUC on A21s test set) and 60% of J8 data (0.86 AUC on J8 test set). At this point, adding any quantity of the other phone’s training data does not significantly alter ROC AUC values in either direction. Within the scope of our experiments, these results indicate that training data from a secondary device is beneficial when primary device data is scarce, but its impact lessens as more primary device data becomes available.

These results and observations are based only on data from the two devices included in this study, so the specific numbers and even the general trend may not apply for different imaging hardware. These are also based only on the specific algorithm approach and architecture described in the main text, and other algorithms may behave differently – for example different data augmentation methods may enable improved device portability.

### Supplement S3. Definition of the target screening population distribution.

### **Rationale and assumptions supporting definition of target population.**

For providers in LMICs, AVE is intended as a primary screening tool or as a triage test for women who are HPV-positive. Our training and testing data from Zambia were enriched with data from patients that underwent LEEP based on an initial screen. This was designed to help our study team collect a sufficient absolute number of images of CIN2+ to train the AVE algorithm. However, it also resulted in an image set with higher severity relative to a typical screening setting. In response, we “rebalanced” our test set, as described in the previous section, which enabled an accurate measure of AVE’s performance in a primary screening or HPV-positive triage setting that would be most typical for the geographies where AVE is intended to be used.

Defining a severity distribution for a primary screening or HPV-positive triage population is not straightforward, as prevalence of different disease states varies between regions and is not always well documented. We therefore made assumptions based on a triangulation of the literature and data from national screening programs in 2020 and 2021.

Specifically, we have assumed:

Among the general population of women (applicable when AVE/VIA is used as a *primary screen*):

- 75% HPV- / no lesions (VIA- or CIN0) based on an assumption of 25% HPV positivity. A 2020 meta-analysis by *Bogale et al* estimates HPV positivity in Sub-Saharan Africa at 24%; 50.8% among WLHIV; and 22.8% among HIV uninfected women.^[1]^ When we apply these rates to the adult female HIV prevalence estimates in the five countries expected to be early adopters of AVE (Malawi, Rwanda, Senegal, Zambia, and Zimbabwe), the estimated HPV prevalence is estimated at 20% - 30%. This aligns with the HPV prevalence seen in programs in these countries.
- 17.5% HPV+ / no lesions (VIA- or CIN0): This is based on program data in settings using HPV testing.
- 4.7% CIN1: Research from India, China, Uganda, and Asia estimates prevalence at 1.3% - 4.7%; we have chosen the upper estimate (from India) to be conservative.^[2],^^[3],^^[4],^^[5],^^[6]^
- 2.3% CIN2/3 prevalence: This is estimated by applying the WHO’s estimate of CIN2/3 prevalence of 10% among WLHIV^[7]^ to the adult female HIV prevalence in presumed AVE early adopter countries, yielding CIN2/3 prevalence estimates of 1% - 3%.
- 0.5% invasive cervical cancer. Some studies have estimated prevalence to be 0.2% - 1.1%.^[8],^^[9],^^[10]^ GLOBOCAN’s estimates are lower – less than 0.1% for all AVE early adopter countries. We use 0.5% as a conservative estimate.

Table S2.1 shows the number and percentage of images in several ‘severity’ levels for the Zambia A21s test set images, based on metadata, as well as the target population prevalence within each severity level. For this target population evaluation, we did not involve test participants who were missing HPV status but kept in participant cases (CIN2+) with missing HPV status since they were already confirmed histology positive. As shown in Table 2 in the main text, 131 test set participants (9.6%) were not associated with an HPV test. Of these, 122 participants were negative and 9 were CIN2+. Overall, our study involved 1024 participants who were HPV-available negative and 216 participants who were CIN2+ (involved in the general population simulation).

*Table S2.1. Proportion of participants in test set: Original test set vs. Target population.*

|  | Severity group | Test set count | Percentage of test set | Target population |
| --- | --- | --- | --- | --- |
| Negative participants (1146) | HPV missing, no lesions (VIA- or CIN0) | 122 | 9.0%  (10.6% of negatives) | Not used. |
|  | HPV-, no lesions (VIA- or CIN0) | 665 | 48.8%  (58.0% of negatives) | 75%  (77.2% of negatives) |
|  | HPV+, no lesions (VIA- or CIN0) | 295 | 21.7%  (25.7% of negatives) | 17.5%  (18% of negatives) |
|  | CIN1 | 64 | 4.7%  (5.6% of negatives) | 4.7%  (4.8% of negatives) |
| Positive participants (216) | CIN2, CIN3 | 174 | 12.8%  (80.6% of positives) | 2.3%  (82.1% of positives) |
|  | Cancer | 42 | 3.1%  (19.4% of positives) | 0.5%  (17.9% of positives) |

**Method**

We simulated the general population via a bootstrapping technique^[11]^ where we sampled the participants from the test dataset with replacement, such that the participant composition of each of the bootstrap samples closely reflected the target population proportions. Participants who were positive for the CIN2, CIN3 and Cancer severity groups were sampled separately from participants who were negative and composed of the other three severity groups in table 1 of the main text. For example, participants who were HPV negative, and thus without lesions, represented 58.0% of participants who were negative in our test set but 77.2% of the population who were negative as shown in table S2.1, so they are upweighted relative to our data. By comparison, cancer cases represented 19.4% of our positive test set, but 17.9% of the population who were positive, as shown in table S2.1, and slightly downweighted as a result. We sampled the dataset 1000 times. Each sample contains two-thirds of the 1024 participants who were HPV-available negative (683) and two-thirds of the 216 participants who were CIN2+ (144). Then, we retrieved all images for the participants in each sample, accounting for repeats (i.e., if a participant was present 3 times in a sample, there were 3 copies of each image).

For each of the 1000 samples, we generated image-level and participant-level sensitivity, specificity, and ROC AUC. For participant-level metrics, we use the “maximum confidence” method outlined in the paper and preserved repeats of participants as needed so that the target population proportions were not altered. From this, we estimated sensitivity, specificity, and AUC from their median values in the 1000 bootstrap samples and produced 95% bootstrap confidence intervals based on the upper and lower bound from the middle 95% of values. Furthermore, we generated metrics for populations who were HPV and HIV positive by simply extracting the corresponding participants from each of the 1000 samples and reproducing the median metrics and 95% confidence intervals.

We generated ROC curves as shown in Figure 3 in the main text by extracting the participant-level scores from the combination of all 1000 bootstrap samples. The combined-sample AUC very closely resembled the bootstrap median AUC numbers, giving us confidence that the curves accurately represented the general population screening.

An alternative bootstrapping sampling technique would be to sample together the participants who are positive and those who are negative to match the whole target population. The challenge with this technique is that the overall population has a strong imbalance between subjects who are positive and those who are negative, whereas the dataset from this study was more balanced (due to some enrollment from referral sites). Sampling participants together to match the target population would result in significant downweighing of the positive classes to the point that each sample would only have a few dozen subjects who are positive, thus artificially inflating the confidence intervals for sensitivity and AUC. As the main metrics sensitivity and specificity do not depend on the ratio between cases and controls, and ROC AUC stems from these metrics calculated at different thresholds, the approach of sampling participants who are positive separately from those who are negative is justified.

To evaluate AVE’s performance when used as a triage test for women who are HPV positive, we used the general population balanced as above and removed all women who are HPV negative. Similarly, we removed all women who are HIV negative to evaluate AVE’s performance on women who are HIV positive.
